# Research priorities for lower limb amputation in patients with vascular disease

**DOI:** 10.1101/2021.05.21.21256746

**Authors:** DC Bosanquet, S Nandhra, KHF Wong, J Long, I Chetter, RJ Hinchliffe, on behalf of The Vascular Society of Great Britain and Ireland Amputation Special Interest Group James-Lind Alliance Priority Setting Partnership

## Abstract

**Introduction:** Lower limb amputation is a life-changing event for patients and can be associated with high mortality and morbidity rates. Research into this critical part of vascular surgery is limited. The Vascular Society of Great Britain and Ireland (VSGBI) in partnership with the James Lind Alliance (JLA) process, aimed to identify and develop key research priorities for amputation.

**Methods:** A modified JLA Priority Setting Partnership was undertaken, encompassing all vascular practice. Two separate Delphi processes to identify research topics were undertaken with healthcare professionals, patients and carers, led by the VSGBI. The priorities were then ranked by the same participants and amalgamated to produce a list for final prioritisation. The final consensus meeting was attended by patients, carers and healthcare professionals from a variety of backgrounds involved in the care of people with amputation. Using a nominal group technique, the top ten research priorities were identified.

**Results:** A total of 481 clinicians submitted 1231 research questions relating to vascular surgery in general. 63 amputation-specific research questions were combined into 5 final clinical questions. 373 patients or carers submitted 582 research questions related to vascular surgery in general. Nine amputation-specific research questions were identified after combining similar questions. Amalgamating both the clinician and patient questions, 12 questions were discussed at the final prioritisation meeting and the top 10 identified. These related to amputation prevention, supporting rehabilitation, improving clinical outcomes following amputation (preventing/treating pain including phantom limb pain and improving wound healing) and research into information provision for patients undergoing amputation.

**Conclusion:** The top 10 research priority areas in vascular amputation provide guidance for researchers, clinicians, and funders on the direction of future research questions that are important to both healthcare professionals and patients.

## Introduction

Over 4000 major lower limb amputations (MLLA) are performed per annum in the UK [1] for end-stage lower limb arterial disease or profound foot sepsis. Amputation is a significant life event for patients and their carers/families. Although supported by recommendations for optimal practice [2], MLLA can be associated with high mortality and complication rates [3,4]. The process of a patient undergoing MLLA is a multi-faceted healthcare challenge, dependent on the complex integration of pre-operative assessment, peri-operative care techniques and postoperative rehabilitation.

Within these care pathways, there are invariably numerous opportunities for interventions to improve clinical and patient-reported outcomes. At present, there is a paucity of high-quality research into MLLA care; for instance, the Cochrane vascular database of 177 systematic reviews contains only two on a topic pertinent to MLLA [5,6]. It is therefore imperative to understand where impactful research, valued by both patients and the healthcare professionals, should be focused, which can inform research funders, health commissioners and policy makers. Arguably, to develop research priorities which are generalizable and of broad value, they should be relevant to the patient and clinician and avoid wasted research efforts [7]. One validated approach is the James-Lind Alliance (JLA) priority setting partnership (PSP). This is a collaborative method of discerning key research questions of multi-disciplinary healthcare professionals and patients/carers with lived experience of the condition [8]. The Vascular Society of Great Britain and Ireland (VSGBI) has worked to define the priorities for vascular research in general from a healthcare professional perspective [9]. This initiative led to the development of nine focused special interest groups (SIGs), one of which is Amputation Surgery. The Amputation Surgery SIG comprises a multi-disciplinary team of clinicians and patients/carers with an interest in furthering research activity in the field of amputation surgery. The aim of this exercise was to create a hierarchical list of important clinical research questions in the field of amputation surgery, using the modified JLA Priority Setting Partnership (PSP), to guide future investigative endeavours.

## Methods

A modified version of the JLA PSP methodology [8] was used to address vascular surgery research priorities in their entirety. The process began with a clinician-led priority setting process, followed by a similar patient-led process. Amputation surgery specific research questions were identified from both processes, duplicates removed and unclear language resolved, before a final priority setting workshop. The Vascular Condition PSP process is summarised in *Figure 1*.

**Figure 1.**
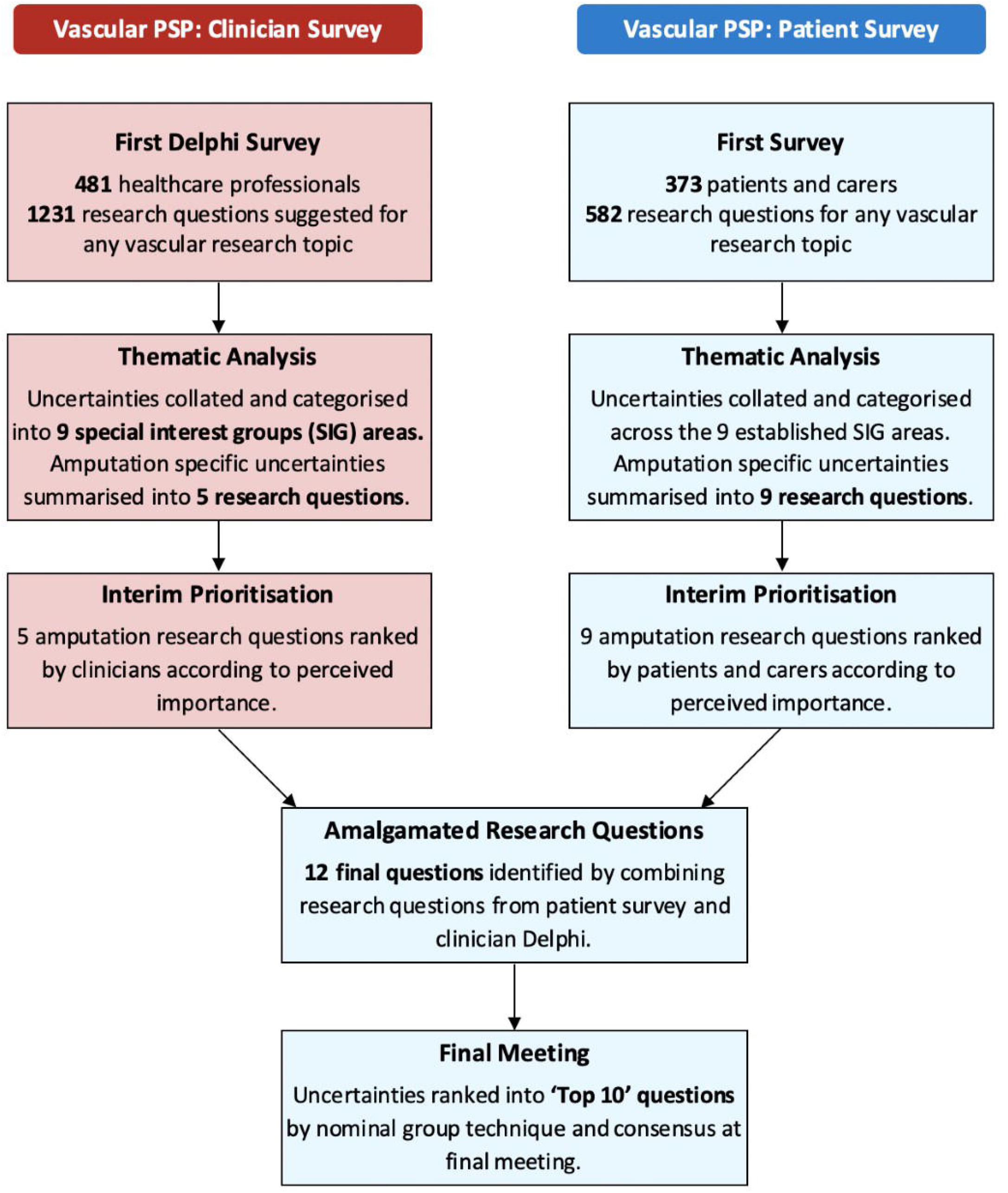
Flow diagram of the Priority Setting Partnership.

Throughout the PSP, the intention was to follow the JLA process as closely as possible. However, initial resource limitation meant that the first survey, gathering research priorities, had to be confined to the clinical community only. The survey gathering questions from patients and supporters followed two years later, when resources permitted. The two sets of questions were analysed, summarised and ranked separately by their respective communities.

### Setting up the Vascular Condition PSP

The Vascular Research Collaborative (VRC) was founded in 2016 and the Vascular Condition PSP was subsequently established in April 2019 [10]. Both were funded by the VSGBI, aiming to develop a national strategy for vascular research and identify research priorities in sub-specialty areas within vascular surgery. The initial outcomes and processes have been published [9].

### Scope of the Amputation Surgery SIG

The Amputation Surgery SIG PSP team comprised two clinical leads (RJH and DB), two surgeons in training (SN and KW), a JLA adviser (TG), one PSP information scientist (JL), one SIG coordinator (BC) and a patient representative (DC), who has bilateral MLLA with experience of amputation research.

The remit of the Amputation Surgery SIG is to support research into the process of a minor or MLLA in adults (18 years or older), including pre-, peri-, and post-operative care, and to develop the top ten research priorities in amputation surgery. For the purpose of the PSP, MLLA was defined as surgical removal of the lower limb above the ankle. Minor amputation was defined as surgical removal of a toe(s) and/or part of the foot. The SIG considered amputations due to peripheral arterial disease, non-healing wounds and/or diabetes related complications within its remit.

### Clinician-led priority setting process

A clinician-led PSP process was completed in 2018 [9], which identified 9 key areas from 45 potential topics using a modified Delphi approach with two rounds of online surveys involving the membership of the VSGBI, Society of Vascular Nurses (SVN), Society for Vascular Technology (SVT) and the Rouleaux Club (vascular surgical trainees). The first round invited any suggestions for research questions in the broad scope of ‘vascular surgery’, which were then collated and categorised into pathological topics and research categories by the steering group. Questions relating to the same fundamental issue were amalgamated into a single question. The questions were recirculated in the second round to the same participants in the second round for priority scoring. These results have been published [9] and are presented here briefly.

### Patient/carer-led research question identification process

The Vascular Condition PSP carried out a consultation to gather potential research questions from vascular patients and carers for approximately 6 months (27 August 2019 to 17 March 2020) via online surveys, paper surveys in outpatient clinics and focus groups. SIG members, UK vascular units (as listed on the National Vascular Registry), charities and patient groups were contacted and asked to distribute (physically and electronically) a survey designed to gather potential research questions in vascular surgery. The affiliated healthcare organisations listed above, the British Society of Interventional Radiology (BSIR) and the British Association of Chartered Physiotherapists in Amputee Rehabilitation (BACPAR) were also asked to circulate the survey. Amputation-specific research questions were identified and similar or duplicate questions merged. Generic questions relating to the overall provision of vascular services (which may have included services related to amputation surgery) were considered outside the remit of the Amputation Surgery SIG and reviewed by the ‘Service’ SIG. Questions were edited by the SIG chairs, with input from the SIG patient representative, to produce a list of easily understood research questions, with no overlap and minimal uncertainty. These minor edits were subsequently ratified by the rest of the SIG team.

### Interim patient/carer led research question prioritisation process

Summarised research questions were put out to interim prioritisation. Questions from each of the SIGs were presented for completion at this point. Patients and carers with experience of amputation(s) were asked to rank questions identified in order of importance to them. This process was undertaken from 5 November 2020 to 27 January 2021. In order for results to be reviewed in time for the final prioritisation workshop, ranking of amputation-related questions was stopped on 9 December 2020.

### Final prioritization workshop

Prior to the prioritisation workshop, the SIG team combined interim prioritised questions with questions from the clinician PSP survey and duplicates were merged. The patient representative was involved in this process and the end-result was again ratified by the Amputation Surgery SIG.

The final prioritisation process was conducted via a virtual online meeting on 25 January 2021. Patient/carer attendees were recruited via direct contact and if they expressed interest in supporting the prioritisation workshop during the research question identification and prioritisation process. Healthcare workers were recruited via direct communication with national bodies (e.g. BACPAR and The Royal College of Occupational Therapy Specialist Section Trauma and Musculoskeletal Health; Prosthetic Amputee Forum; RCOTSST&MSH PAR) and via direct links with members of the SIG team.

The workshop was led by three advisers skilled in the JLA process. Members of the Amputation Surgery SIG provided general support, but had no influence over the process of priority setting. A nominal group technique was used to reach the final top 10 research priorities. Workshop attendees were asked to review the final research questions prior to attending the meeting and rank them in order of importance. After an overview of the JLA process, attendees were divided into three ‘breakout’ groups, each comprising an equal mix of patients, carers and healthcare professionals. The ordering of research questions was discussed three times. In the first breakout group, each participant presented their ‘top three’ and ‘bottom three’ of the shortlisted questions. In the second round, the same groups discussed how to pool these individual rankings into a single priority listing (numbered 1-12). The priority listings from the three groups were collated to generate an interim ranking of the research questions. Finally, the attendees were allocated to different groups for a third round of breakout discussion, to discuss the finer details of the order of the interim ranking. The results of each group’s rankings were again collated and summated, creating a final list. The finalised list of top 10 research priorities was presented to participants in a final session to facilitate discussion of overall acceptability. Members of the SIG PSP team observed all sessions (muted with cameras off) and noted key points arising from the discussion.

## Results

### Results from the clinician led research question identification and prioritisation

Some 481 healthcare professionals involved in the care of vascular patients engaged with the vascular surgery PSP, suggesting a total of 1231 research questions [9]. 63 amputation specific research questions were reported. After combining similar research questions, a final list of five important amputation research questions was identified and redistributed to clinicians for scoring regarding importance. Questions were ranked according to clinicians’ scores. The resulting clinicians’ research questions ordered by importance, along with a mean score, are given in Table 1.

**Table 1.**
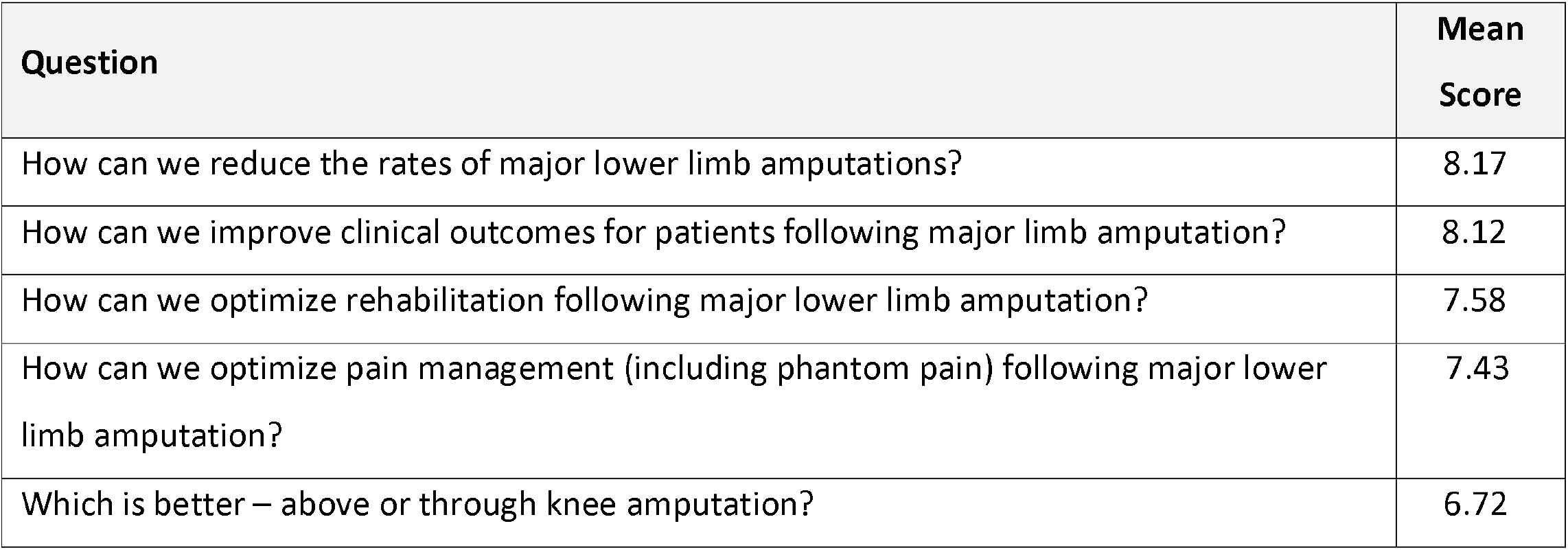
Questions arising from the clinician question identification and prioritisation process, with their mean score obtained from ranking.

### Patient/carer led research question identification and prioritisation

373 vascular patients/carers suggested a total of 582 research questions, relating to vascular surgery in general. 14 were specific to amputation surgery. After combining overlapping questions, a total of nine research questions were confirmed and redistributed to patients / carers for scoring regarding importance. 36 patients/carers engaged with the scoring process. The resulting patient research questions, ordered by importance and along with a mean score, are given in Table 2.

**Table 2.** Questions arising from the patient/carer question identification and prioritisation process, with their mean score obtained from ranking.

| Question | Mean Score |
| --- | --- |
| In a person who has undergone amputation, how do you reduce the chances of amputation in the other limb? | 4.69 |
| What are the best ways to support rehabilitation following amputation? | 4.57 |
| How do we optimise prosthetic limb use following amputation? | 4.55 |
| When is it appropriate to perform a major amputation? | 4.39 |
| What are the best ways to prevent or treat pain after amputation? | 4.36 |
| In a person who has undergone a minor amputation in the foot, how are the chances of a subsequent major lower limb amputation above the ankle reduced? | 4.34 |
| How do you improve healing of the amputated stump? | 4.28 |
| What are the best mobility aids following amputation? | 4.26 |
| How do we improve the information provided to patients undergoing amputation? | 4.17 |

### Final prioritization workshop

Prior to the workshop, the Amputation Surgery SIG team pooled clinician and patient/carer research questions, resulting in a final list of 12 questions, detailed in Table 3. In order to reduce risk of bias, these questions were randomly ordered and each assigned a letter (rather than a number).

**Table 3.**
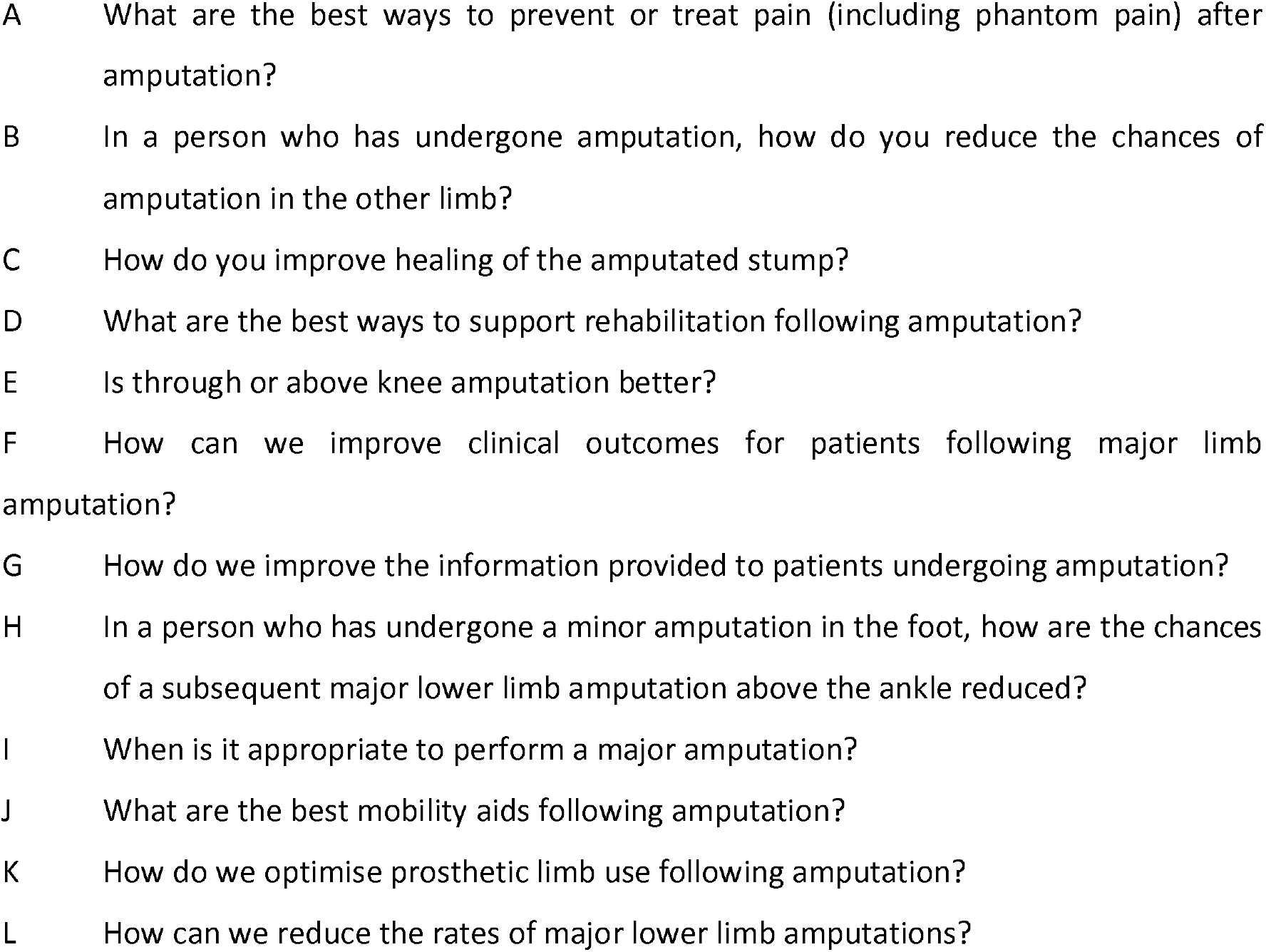
Collated research questions, listed by letter (not number) for review by participants prior to the prioritisation meeting.

The final prioritisation workshop was attended by 10 patients/carers and 12 healthcare professionals, with an additional 8 observers. The prioritisation process resulted in the final top ten research priority list (Table 4). The questions are ordered according to importance as determined by the workshop. The last three priorities all scored the same and are therefore ranked equal. There was general consensus amongst the participants that the list accurately and comprehensively reflected well the discussions and viewpoints which occurred in the breakout groups.

**Table 4.**
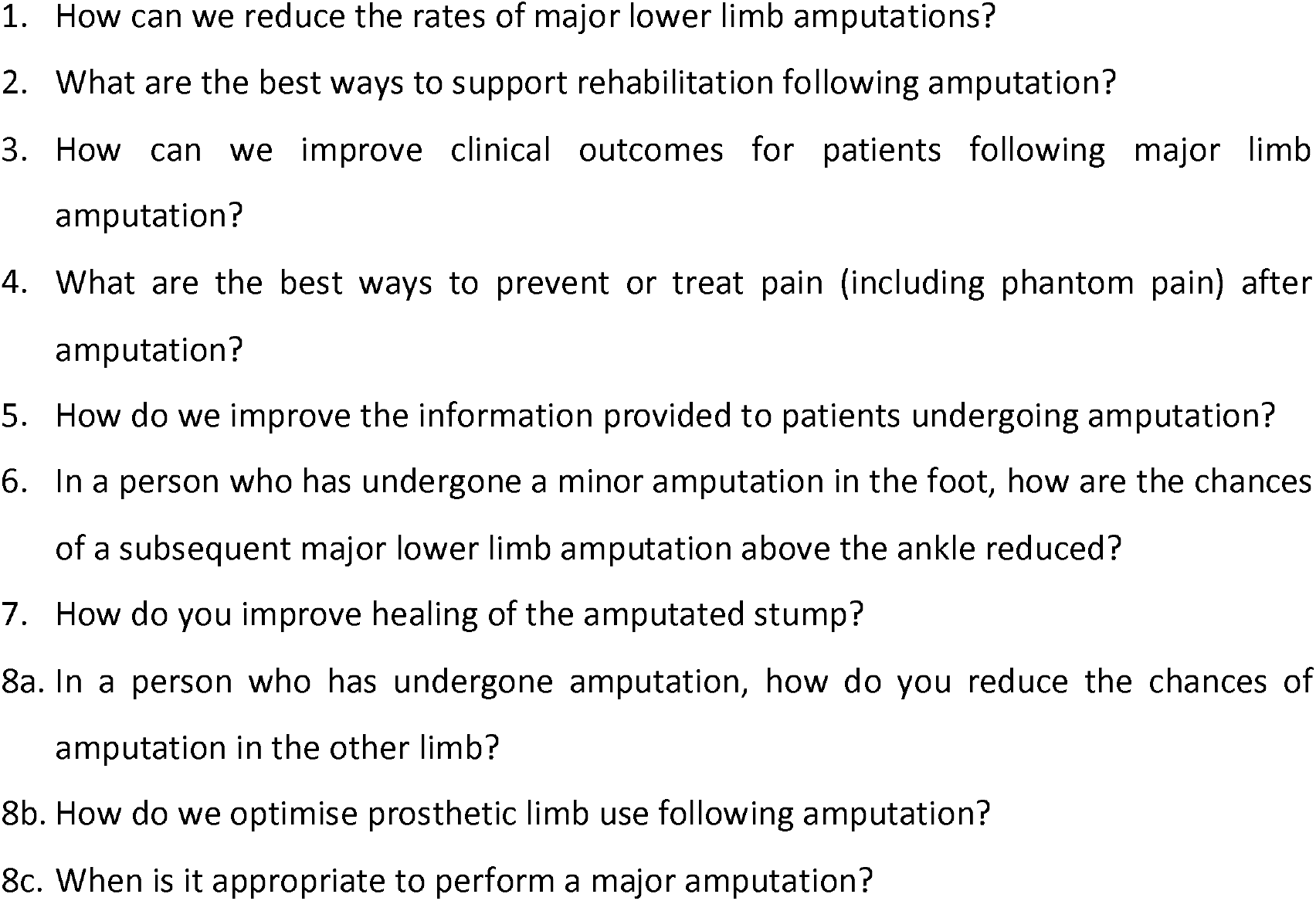
Final ordered list of ‘top ten’ priority research questions in amputation surgery. Note the last three of the ‘top ten’ scored the same, and are considered equal 8^th^ priority.

A number of key points were noted during the discussion. There was clearly a difference between participants who thought amputation prevention was paramount (priorities 1, 6 and 8a) and those who felt improving outcomes following amputation was paramount (priorities 2, 3, 4, 5, 7, 8b and 8c). Many participants individually remarked that there was significant overlap between research questions. For example, priority 1, 6, and 8a all pertained to amputation rate reduction. It was also noted that priority 3 (improving clinical outcomes following amputation) could encompass some of the other research questions, such as improving healing (priority 7) and pain (priority 4) outcomes. Priority 2 (what are the best ways to support rehabilitation following amputation) could encompass priority 8b (How do we optimise prosthetic limb use following amputation) and priority 12 (What are the best mobility aids following amputation).

The two research priorities which did not make the top-10 were priority number 11 (“Is through or above knee amputation better?”) and 12 (“What are the best mobility aids following amputation?”). It was noted that the low ranking of priority 11 (through knee versus above knee amputation) may have been influenced by through-knee amputations being less commonly performed in the UK. Furthermore, there were no patients with a through knee amputation in the workshop. Participants expressed that the lack of awareness and experience within this cohort (particularly from the patient representatives present) could have led to a perceived lower importance of this research question.

## Discussion

### Summary

Using a modified JLA methodology in priority setting, we identified key research priorities in amputation surgery. A two-round Delphi process covering all aspects of vascular surgery care identified 5 amputation research questions from clinicians, which were pooled with questions raised by patients and carers to produce 12 questions for final prioritisation. Following discussion with patients, carers and healthcare professionals, a top 10 list of clinical research questions in amputation was produced by consensus.

### Strengths and Limitations

The strengths of this study include the use of a well-established systematic and transparent process to identify research priorities of patients and healthcare professionals across the UK, under supervision from a steering group and experienced JLA advisors. The priority setting process included a variety of stakeholders, to provide a broad view of unanswered questions. Facilitation by JLA advisors ensured that all parties contributed actively to discussion.

There are several limitations to consider in this PSP. Firstly, due to the survey-based nature of the process, there is a potential for responder bias for both clinicians and patients, which may not be representative of all patients with amputations and healthcare professionals involved in their care. We attempted to include patients from a wide range of geographical, socio-economic and health literacy backgrounds, as well as healthcare professionals who may interact with patients differently (carers, nurses, doctors, AHP’s). However, not all were available for the final prioritisation process. In particular, there is potential bias involved when considering the research question of comparing through-knee versus above knee amputations: the lack of awareness, experience and representation in this cohort may have impacted on the relative prioritisation of this topic.

The PSP was conducted using a modified approach to conventional JLA priority setting methodology, to capture the broad scope of questions in vascular care. The key modification was the disconnect between the patients and healthcare professional research question identification and prioritisation, which were only pooled later. Typically both groups undertake a single identification and prioritisation process. Due to the inherent subjective process of PSP, prioritisation may have been biased by initial survey approaches. It is not possible to assess whether the top 10 research priorities may have differed, had all questions been analysed, summarised and ranked by all participants. However, it is clear that the chosen top 10 priorities included those of specific importance to patients/carers, to clinicians and to both groups. Finally, the overlap in research questions occasionally made the ranking process more difficult; often participants grouped certain questions together or ranked them higher if they encompassed parts of other topics. Missing research topics identified from the 12 research questions included mental wellbeing and psychological support. These questions do feature in the Service SIG however, encompassing priorities spanning the entire scope of vascular surgery. It was also noted by participants that there was no podiatry representation, recognised as an important stakeholder group

### Implications for future research

Defining a specific top 10 research questions provides an invaluable starting point for future research in amputation surgery. The top 10 research priorities will guide researchers and funders to the most important research questions for both healthcare professionals and patients. Specific research strategy will be decided upon on by further evaluation of individual research questions. Amputation surgery research in the United Kingdom and the wider global amputation community is likely to be guided by this work for many years to come. It is important to recognise that all priorities discussed were considered of value: priorities 11 and 12 remain important areas for future research. It is expected the Amputation Surgery SIG will select individual research priorities, with the aim of specifically developing ongoing research strategy. The overall aim of the Amputation Surgery SIG is to develop a national research group for amputation surgery, with research and amputation experts from around the country, supported by national bodies, such as the VSGBI and the Vascular and Endovascular Research Network (VERN).

## Data Availability

Derived data supporting the findings of this study are available from the corresponding author on reasonable request.

## Acknowledgements

The authors would like to thank the organisation and charities that supported the Amputation SIG JLA process:

BAPO - British Association of Prosthetists & Orthotists

BACPAR - British Association of Chartered Physiotherapists in Amputee Rehabilitation BSIR - British Society of Interventional Radiology

SVN - Society of Vascular Nurses

SVT - Society for Vascular Technology The Limbless Association

The Rouleaux Club

The Royal College of Occupational Therapy Specialist Section Trauma and Musculoskeletal Health; Prosthetic Amputee Forum; RCOTSST&MSH PAR

We would also like to thank all patients, carers and clinicians who responded to the survey(s).

